# Paradoxical relief after seizures: a diagnostic signal distinguishing functional/dissociative from epileptic seizures

**DOI:** 10.64898/2026.08.27.26360607

**Authors:** Ajay Masharani, Akihiro Koreki, Mario Marcelo, Kamiar Shalfrooshan, Ma-an Diamos, Catherine Santucci, Kiran Pillai, Dot Bindman, Suzanne O’Sullivan, Fergus Rugg Gunn, Meneka Sidhu, Mahinda Yogarajah

## Abstract

**Objective:** To determine whether paradoxical relief, feeling unusually better after a seizure compared to before it, is more common after functional/dissociative seizures (FDS) than epileptic seizures (ES), quantify its diagnostic accuracy, and explore its relationship with preictal symptoms.

**Methods:** Consecutive patients admitted to a tertiary epilepsy unit for prolonged in-patient EEG monitoring underwent a structured clinical interview on admission, before final multidisciplinary diagnostic classification. Preictal dissociative and autonomic/somatic symptom burden was assessed using items adapted from established questionnaires. Diagnostic classification incorporated clinical history, seizure semiology, video-electroencephalography findings, and collateral information. Patients with dual or indeterminate diagnoses were excluded. Associations with paradoxical relief were examined using logistic regression, followed by an exploratory mediation analysis.

**Results:** Of 176 patients assessed, 66 with FDS and 65 with ES were included. Paradoxical relief was reported by 46/66 patients with FDS (69.7%) and 10/65 with ES (15.4%; unadjusted odds ratio [OR] 12.65, 95% confidence interval [CI] 5.57–31.09). As a diagnostic signal for FDS, paradoxical relief had 69.7% sensitivity (95% CI 57.1–80.4), 84.6% specificity (CI 73.5–92.4), a positive likelihood ratio of 4.53 (2.51–8.19), and a negative likelihood ratio of 0.36 (0.24–0.52). FDS diagnosis remained independently associated with paradoxical relief after adjustment (OR 10.59, 95% CI 3.42–38.06). In a parallel mediation analysis, dissociative symptom burden showed a significant indirect effect, accounting for 19.5% of the association between diagnostic group and relief, whereas the indirect effect through somatic/autonomic symptom burden was not significant.

**Significance:** Paradoxical relief is substantially more common after FDS than ES and may provide a simple, clinically useful diagnostic signal. Its absence does not exclude FDS, and the finding requires external validation. The association with dissociative symptoms is exploratory and supports prospective investigation of whether relief reflects transient resolution of a disturbed, disembodied preictal state.

## Introduction

Functional / dissociative seizures (FDS) are paroxysmal events that resemble epileptic seizures (ES) but are not associated with epileptogenic discharges on an electroencephalogram (EEG)^1^. Individuals with FDS are at least as common as those with multiple sclerosis^2–4^ and have elevated morbidity^5^ and mortality rates^6^, and health costs^7^ equivalent to those individuals with epileptic seizures. Part of the reason for this is that many individuals with FDS are misdiagnosed for several years and treated as ES exposing them to iatrogenic harm^8^. While the gold standard diagnosis of FDS is based on video telemetry EEG (VT-EEG), or video of FDS with typical semiological characteristics but no associated epileptic discharges, in practice this investigation is not widely available^1^. In diagnostic practice, clinicians rely on the objective characteristics of a seizure, that is, the witness description or videos of the seizures^9^. While these can be useful, in isolation, no single observation has sufficient sensitivity and specificity to be used to help clinicians reliably to diagnose FDS from ES. In addition, eyewitness descriptions are unreliable and this should be borne in mind when using them^10^. While observers often recall the general features of a seizure, they may miss or misreport subtle details because of the stressful nature of their circumstances. Videos, particularly from accessible smartphones, can be more useful in distinguishing FDS from ES, and other events. However, the utility of videos is limited by their quality^11,12^. The best quality videos show the patient’s entire body and face with an unobstructed view before, during, and after the seizure^11^. In addition, accurate interpretation requires expertise, and studies have demonstrated seizure type recognition varies by the reviewer’s experience level of dealing with seizures^13,14^. Videos alone therefore can be insufficient for a confident diagnosis and must be evaluated in an expert manner alongside clinical history and other data.

An underused source of diagnostic information is the subjective experience of the event itself. Large studies based on interviews with individuals about their seizure experience have shown that individuals with FDS typically experience somatic / autonomic and/or dissociative symptoms before and during their seizures. The presence of these symptoms alone can yield sensitivity and specificity rates of up to 80%^15^. While this can also be present in some ES it is much less common and typically associated with temporal lobe seizures^16^. Prominent mechanistic models of FDS propose that the seizure itself is a response to these preceding symptoms^17–19^.

Another potentially useful component of seizure experience is the postictal state. In a case series of 11 individuals with FDS, they experienced a sensation of post-ictal relief linked directly to the overwhelming nature of the symptoms immediately preceding the seizure^20^. In a larger study of 100 individuals with FDS 67% reported experiencing some relief after their seizures, with 32% reporting it was always or frequently present^21^. However, the presence of post-ictal relief, or ‘paradoxical relief’ as we will refer to it below, has never been directly compared between ES and FDS cohorts to quantify its diagnostic utility.

## Objective

To compare the prevalence of post-ictal paradoxical relief in FDS versus ES and evaluate its diagnostic utility.

## Methods

### Design and setting

Consecutive admissions to the Chalfont Centre for Epilepsy undergoing prolonged in-patient EEG monitoring between 18/12/23 and 25/11/24 were considered. This study used routinely collected prolonged EEG and associated clinical information from routine clinical care at the Chalfont assessment unit. Retrospective analysis of these deidentified data was approved as part of a wider study of clinical EEG data by NHS Research Ethics Committee (IRAS References 305776), in conjunction with the University College London Hospitals/University College London (UCL/UCLH) Joint Research Office. The evaluation of the clinical utility of the structured peri-ictal history was additionally registered with the National Hospital for Neurology and Neurosurgery Research Audit and Clinical Governance Committee as a service evaluation (120-202425-SE).

### Participants

All participants underwent prolonged inpatient EEG monitoring, and diagnostic classification was determined by multidisciplinary review incorporating clinical history, seizure videos, video-EEG findings, and available collateral information. FDS were diagnosed in accordance with ILAE staged diagnostic guidance and were included where the diagnosis met documented or clinically established levels of certainty. Documented FDS required capture of a typical event on video-EEG without associated ictal epileptiform activity. Clinically established diagnoses were based on expert multidisciplinary assessment of typical history, witnessed semiology, and/or seizure videos judged characteristic of functional/dissociative seizures. Epilepsy diagnoses were based on captured electroclinical or electrographic epileptic seizures during prolonged EEG monitoring, or on interictal epileptiform abnormalities supported by clinical history, witnessed semiology and/or seizure videos judged consistent with epilepsy. Those participants with an intellectual disability that prevented accurate reporting of their seizure experiences were excluded.

### Ascertainment of symptoms

On admission to the epilepsy unit, part of the routine clinical assessment of patients includes a structured clinical interview by the admitting physician. This includes documentation of symptoms before, during, and after events. Existing literature suggests that pre-ictal autonomic/somatic and dissociative symptoms, together with post-ictal relief, might provide clinically relevant information when distinguishing functional/dissociative from epileptic seizures. These domains are therefore incorporated into the unit’s standard structured admission proforma to support clinical characterisation and multidisciplinary diagnostic assessment. The present study retrospectively analysed the resulting clinical data. Given that the assessment is carried out on admission, at the time of the interview, the final multidisciplinary diagnosis had not yet been made, although the admitting physician is not blinded to all clinical information available at admission.

Interviews use open questions (“Do you get any warning symptoms before your seizures?”) followed by standardised closed questions covering pre-ictal somatic/autonomic and dissociative features (see Appendix 1). Clinical studies have demonstrated that the efficient acquisition of pre-ictal seizure information needs both open and closed questions^22^. These questions comprise items selected and adapted from validated and published questionnaires, namely the Cambridge Depersonalisation Scale^23^ and the Seizure Symptoms Questionnaire^17^. Because item selection, wording, timeframe and response format were modified, the resulting domain scores were treated as study-specific exploratory measures of pre-ictal symptom burden. Individuals rated the frequency with which they experience such symptoms prior to their seizures on a scale from 0 (not at all) to 4 (always). Scores were then summed for each symptom burden (dissociative and autonomic / somatic). Interviews also used open questions regarding post-ictal symptoms (“How do you feel after a seizure?”) followed by the closed question: “Do you in a strange or unusual way, feel better after your seizure compared to before it?”. Response options are recorded as yes/no, and if yes, where possible individuals are asked to elaborate on why they feel better. Individuals who answered “yes” to the standardised paradoxical-relief question were classified as positive unless their accompanying clinical description indicated only simple relief that the seizure had ended or that uncertainty about its occurrence had resolved. Where no further elaboration was available, the affirmative response was retained, reflecting the wording of the question, which specified feeling better in a “strange or unusual way.” Although the presence of paradoxical relief was not used as an established diagnostic criterion, responses were recorded within the clinical assessment and may have been accessible to the multidisciplinary team.

All patients undergo (or have previously undergone) formal interview-based neuro-psychiatric assessments. The presence of depression/anxiety based on these assessments was also recorded.

### Statistical Analysis

Group differences were examined using Welch’s t-test for continuous variables and the chi-square test for categorical variables, as appropriate. To compare the prevalence of post-ictal “paradoxical relief” in FDS versus ES, a logistic regression model was performed with paradoxical relief (presence vs. absence) as the dependent variable and group, age, sex, age at onset, somatic symptom burden, and dissociative symptom burden as independent variables.

To explore whether the association between diagnostic group and paradoxical relief was statistically accounted for by preictal symptom burden, a parallel multiple mediation analysis was conducted with diagnostic group as the independent variable, dissociative and somatic/autonomic symptom burdens as parallel mediators, and paradoxical relief as the outcome. To account for the overlap between the two symptom domains, the residual covariance between the two mediators was freely estimated. Finally, to evaluate diagnostic usefulness, an additional logistic regression model was performed with diagnostic group as the dependent variable and paradoxical relief (presence vs. absence), age, sex, age at onset, somatic symptom burden, and dissociative symptom burden as independent variables.

## Results

A total of 176 patients were included. Of these, 18 patients could not be definitively diagnosed, and one patient was diagnosed with migraine, and these patients were therefore excluded. To ensure a clear distinction between FDS and ES, 26 patients with a dual diagnosis of both conditions were also excluded. Ultimately, 66 patients with FDS and 65 patients with ES were included in the analysis. Compared with the ES group, the FDS group was younger and had a higher proportion of females. The FDS group also had a higher proportion of habitual seizures captured during video telemetry (VT) and an older age at onset.

Paradoxical relief was reported significantly more frequently in the FDS group than in the ES group (46/66 vs. 10/65, p < 0.001, unadjusted odds ratio [OR] 12.65, 95% confidence interval [CI] 5.57–31.09). In addition, both dissociative and somatic symptoms were more frequently reported in the FDS group than in the ES group (Table 1).

**Table 1:** Characteristics of patients and their seizures.

|  | FDS (n=66) | ES (n=65) | P-values |
| --- | --- | --- | --- |
| **Patients' background** |  |  |  |
| Age, mean ± SD, years | 36.6 ± 14.8 | 42.5 ± 14.1 | **0.022** |
| Sex, Female (%) | 51 (77) | 38 (58) | **0.034** |
| Age of onset, mean ± SD, years | 26.2 ± 15.3 | 17.0 ± 13.7 | **<0.001** |
| Habitual event captured during VT EEG, N (%) | 51 (77) | 23 (35) | **<0.001** |
| Anxiety/depression comorbidity, N (%) | 47 (71) | 41 (63) | 0.421 |
| **Paradoxical relief and symptoms prior to their seizures** |  |  |  |
| Paradoxical relief, N (%) | 46 (70) | 10 (15) | **<0.001** |
| Dissociation, mean ± SD | 9.3 ± 9.6 | 3.8 ± 6.9 | **<0.001** |
| Somatic symptoms, mean ± SD | 13.2 ± 8.7 | 4.9 ± 6.0 | **<0.001** |

ES: epileptic seizure, FDS: functional dissociative seizure, SD: standard deviation, VT: Video telemetry,

To test whether the primary finding persisted in patients with the highest diagnostic certainty, we performed a sensitivity analysis restricted to those in whom a habitual event was captured during VT-EEG. In this subgroup, paradoxical relief remained more common in FDS than ES: 35/51 patients with FDS reported paradoxical relief compared with 1/23 patients with ES (OR 48.12, 95% CI 8.90–899.71). Although the confidence interval was wide, the direction and magnitude of the association supported the robustness of the primary finding.

The logistic regression model demonstrated that group was significantly associated with paradoxical relief (odds ratio [OR] = 10.59, 95% confidence interval [CI] = 3.42 to 38.06, p < 0.001). Dissociative symptom burden was also a significant predictor (OR = 1.13 per unit increase, 95% CI = 1.04 to 1.23, p = 0.004). Age, sex, age at onset, and somatic symptom burden were not significant predictors.

In a parallel multiple mediation analysis, there was a significant indirect effect of diagnostic group on paradoxical relief through dissociative symptom burden (standardised indirect effect = 0.12, 95% CI 0.03–0.21, p = 0.010). In contrast, the indirect effect through somatic/autonomic symptom burden was not significant (standardised indirect effect = 0.06, 95% CI −0.05 to 0.17, p = 0.274). However, the difference between these indirect effects was not statistically significant (p = 0.506). The total effect of diagnostic group on paradoxical relief was significant (β = 0.61, p < 0.001) and was attenuated but remained significant after adjustment for both mediators (direct effect β = 0.43, p < 0.001), consistent with partial mediation. Approximately 19.5% of the total effect was mediated through dissociative symptom burden (figure 1). To account for overlap between preictal dissociative and autonomic/somatic symptom burden, the residual covariance between the two mediators was freely estimated. The standardised residual covariance was 0.58, indicating substantial residual association between symptom domains after accounting for diagnostic group.

**Figure 1.**
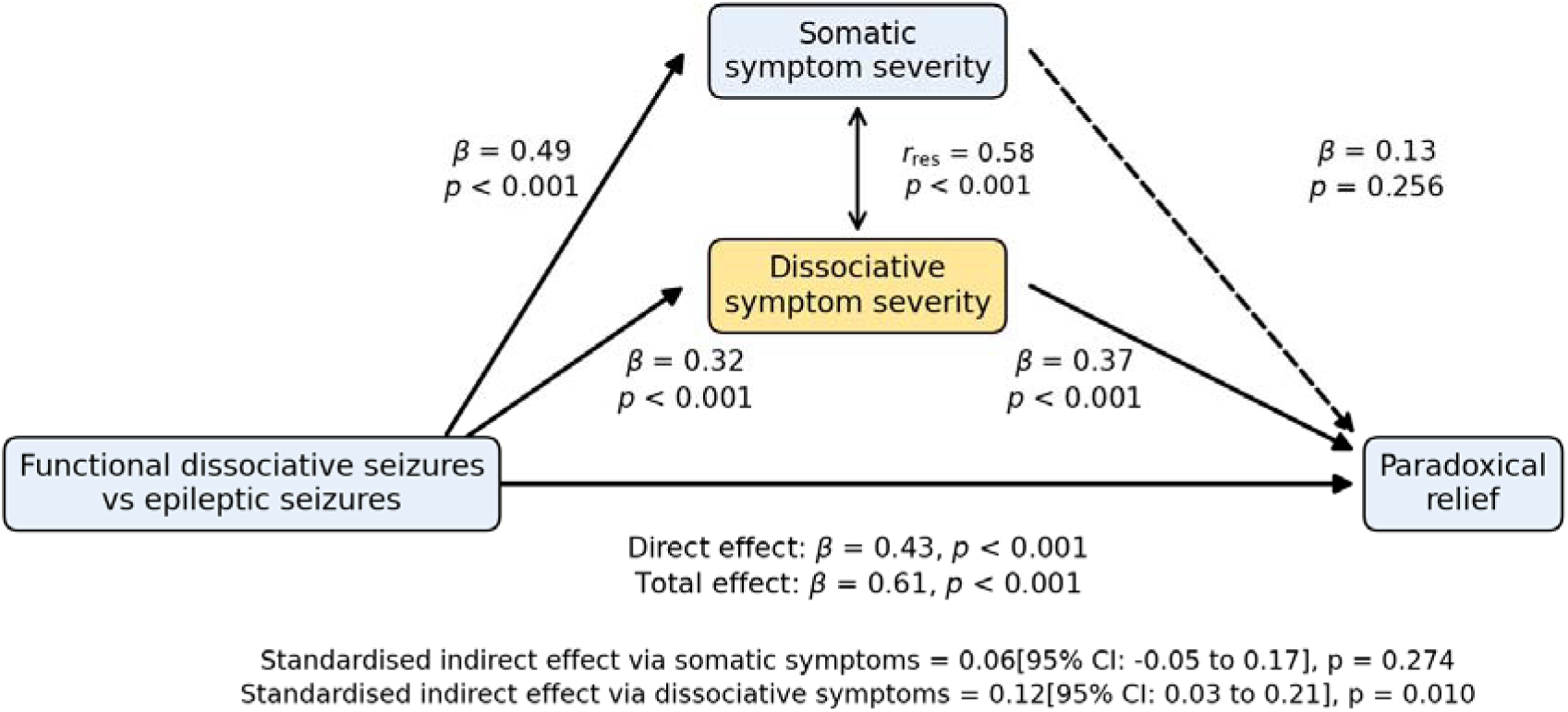
A parallel mediation model A parallel mediation model was used to examine the association between diagnostic group and paradoxical relief through somatic/autonomic and dissociative symptom domains. The indirect effect through dissociative symptom burden was significant, whereas the indirect effect through somatic/autonomic symptom burden was not. The total effect of diagnostic group on paradoxical relief was significant and was attenuated but remained significant after adjustment for both mediators, consistent with partial mediation. r_res_ represents the residual correlation, i.e. the standardised residual covariance. CI, confidence interval.

In the diagnostic model, paradoxical relief significantly predicted FDS (OR = 10.15, 95% CI = 3.10–39.11, p < 0.001). Younger age (p = 0.001), older age at onset (p < 0.001), and higher somatic symptom burden (p = 0.003) were also significant predictors. By contrast, dissociative symptom burden was not significant (p = 0.378), suggesting that its diagnostic contribution may have been partly captured by paradoxical relief. Paradoxical relief alone showed a sensitivity of 0.697 (57.1-80.4) and a specificity of 0.846 (73.5-92.4) for the diagnosis of FDS with a positive likelihood ratio of 4.53 (2.51–8.19), and a negative likelihood ratio of 0.36 (0.24–0.52) respectively.

Among patients classified as experiencing paradoxical relief, descriptions suggested that this was not simply relief that the seizure had ended, but often involved a perceived shift in bodily state, energy, or embodied coherence. Illustrative examples included: “It is as if I feel earthed or fused again … whole again”; “I feel whole again … like I am back in my body”; “Sometimes it feels like it needed to happen to get my energy back”; “This strange build up and strange taste has gone … I am kind of glad it has happened”; and “I feel like I have been energised afterwards…”.

## Discussion

Paradoxical post-ictal relief was substantially more common after FDS than ES, and when considered alone demonstrated moderate sensitivity and high specificity for functional seizures. In an exploratory multivariable analysis, FDS remained independently associated with paradoxical relief. In a separate diagnostic model, paradoxical relief and pre-ictal autonomic/somatic symptom burden independently predicted FDS. In contrast, while pre-ictal dissociative symptoms independently predicted paradoxical relief and partially mediated the relationship between seizure type and relief, dissociation itself did not independently contribute to diagnostic classification once paradoxical relief was included in the model. Together, these findings suggest that dissociative symptoms may contribute mechanistically to the generation of paradoxical relief, whereas paradoxical relief itself may represent a more clinically accessible and diagnostically useful downstream experiential manifestation of these processes. More broadly, these results support the view that subjective peri-ictal experiences, particularly changes occurring after seizures, may contain diagnostically and mechanistically relevant information that is underutilised in current approaches to seizure assessment.

### Paradoxical relief

Seventy percent of patients with functional seizures reported paradoxical relief. This is in keeping with one of the earliest studies to explore post seizure relief which reported that 67% of patients with functional seizures experienced relief after their seizures^21^. However, this study is the first to directly compare the presence of paradoxical relief in patients with epileptic seizures, showing that it was present in only 15% of those patients. Patients with functional seizures were 13 times more likely to report the presence of paradoxical relief alone, and using exploration of this subjective experience alone, achieved a diagnostic sensitivity and specificity of 0.70 and 0.85 respectively. In combination with other information, it may therefore provide useful diagnostic information (figure 2).

**Figure 2.**
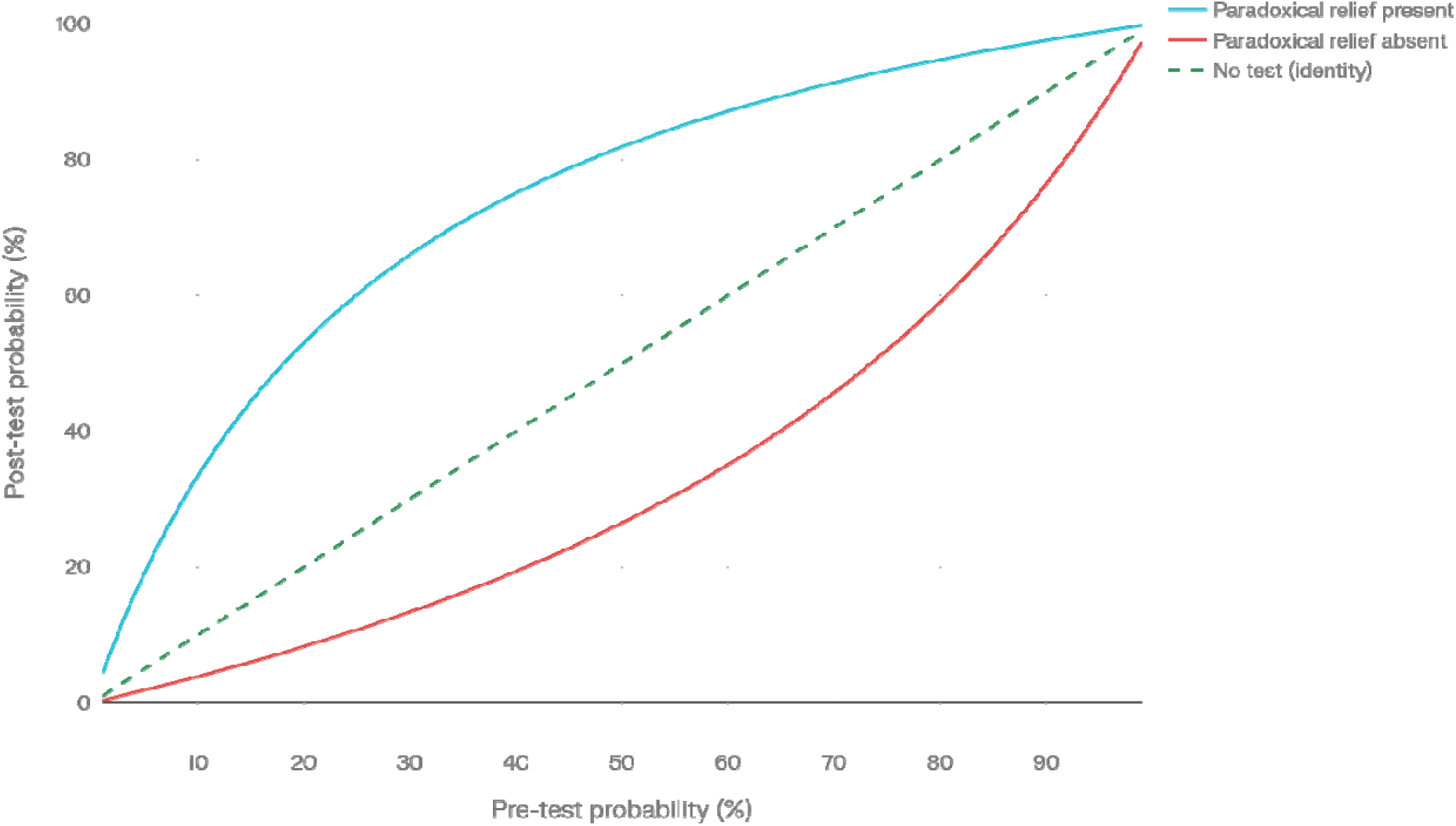
Effect of paradoxical relief on the probability of functional/dissociative seizures. Pre-test probability of functional seizures (x-axis) is plotted against post-test probability (y-axis) when paradoxical relief is present (solid line) or absent (dashed line), compared with the diagonal line indicating no change in probability. Curves are calculated from the observed sensitivity/specificity and positive/negative likelihood ratios of paradoxical relief for functional seizures. The figure illustrates how this single history feature can substantially increase or decrease the post-test probability of a functional seizure across a range of plausible pre-test probabilities.

Although paradoxical relief was far more common after functional seizures, it was not completely absent in those individuals with epileptic seizures. It is possible that the presence of pre-ictal dysphoria which is then improved by the epileptic seizure could explain this finding^24–26^. Although anticipatory anxiety of seizures may be present in up to 50% of drug refractory epilepsy patients, around 25% also have post-ictal anxiety^27^ making this a less likely explanation for our findings in those with epilepsy. Furthermore, we attempted to ensure that individuals describing simple relief related to anticipatory anxiety or concern about the occurrence of the seizure itself were not included.

### Mediation by Pre-Ictal Symptom Burden

The parallel mediation analysis suggested that the association between FDS and paradoxical relief was statistically accounted for in part by dissociative symptom burden, but not significantly by somatic/autonomic symptom burden (figure 1). This supports the possibility that paradoxical relief is more closely related to transient resolution of a dissociated preictal state than to reduction of bodily arousal alone. However, because the indirect effects through dissociative and somatic/autonomic symptoms did not differ significantly, this apparent specificity should be regarded as preliminary. Interestingly, while dissociative symptoms predicted paradoxical relief, they did not independently improve diagnostic classification once paradoxical relief itself was included in the diagnostic model. This suggests that paradoxical relief may represent a more clinically accessible downstream experiential manifestation of these underlying processes.

One interpretation of the mediation findings is that paradoxical relief may emerge not simply from reduction in bodily arousal after seizures as has previously been proposed^19,28^, but from transient resolution or reorganisation of a disturbed (dissociated) pre-ictal embodied state. Within this framework dissociative symptoms may represent a higher-order disruption in embodied self-representation, in which the body no longer feels fully coherent, present, or integrated. Functional seizures may then act as maladaptive, allostatic responses that transiently restore bodily salience, coherence, or embodied presence. This would result in the post-ictal experiences of clarity, energy, or relief described by many patients, despite the distressing nature of the events themselves. This interpretation is consistent with our previous published model of functional seizure semiology, in which altered interoceptive (bodily) attention and disturbed embodiment were proposed as central mechanisms underlying functional seizures^18,29^.

### Dissociation, Autonomic symptoms and Diagnosis

When assessing the utility of pre- and post-ictal symptoms at distinguishing between epileptic and functional seizures, it was clear that paradoxical relief and pre-ictal autonomic symptoms, rather than dissociative symptoms were diagnostically useful. While large studies using self-report questionnaires^30^ or semi-structured interviews^15^ have shown that autonomic and dissociative symptoms can help distinguish between functional and epileptic seizure, they have tended to focus on the presence of such symptoms during and/or after seizures. In those studies using self-report-questionnaires^31^ or semi-structured interviews^32^ to study symptoms before functional or epileptic seizures, autonomic/somatic symptoms appear to be more useful diagnostically than dissociative symptoms. This is not surprising in many respects, given the significant overlap in dissociative symptom experience in both epileptic and functional seizures^16,33^. While dissociative symptoms are mechanistically relevant and partially explain the emergence of paradoxical relief, they are not independently diagnostic once relief is accounted for. This likely reflects the fact that dissociation occurs across both epileptic and functional seizures. In contrast, paradoxical relief may represent a more specific downstream experiential manifestation of these processes and therefore provides greater diagnostic discrimination.

### Therapeutic implications

These findings raise the possibility that, in some patients, transient resolution of a distressing pre-ictal state could negatively reinforce the seizure response. If confirmed, these findings would suggest that therapeutic approaches may benefit from targeting both extinction of the conditioned seizure response and development of alternative strategies that reduce preictal dissociative intensity, improve tolerance of dissociative experiences, or teach alternative coping strategies.

### Limitations

This study has several limitations. First, it was conducted in a single tertiary epilepsy assessment unit using prolonged in-patient EEG monitoring, and the findings may not generalise to less selected outpatient populations or to services without specialist seizure expertise. Second, because pre-ictal symptom reports and paradoxical relief were recorded as part of routine clinical assessment and may have been accessible to the MDT, incorporation bias cannot be fully excluded. However, the association between FDS and paradoxical relief persisted in a sensitivity analysis restricted to ES and FDS patients with habitual events captured during VT-EEG. Third, paradoxical relief and pre-ictal symptoms were ascertained using routine clinical interviews and items adapted from established questionnaires respectively. However, neither have been independently validated introducing potential recall, interviewer, and classification bias. The symptom-domain and mediation analyses should therefore be regarded as exploratory and hypothesis-generating, requiring replication using prospectively validated peri-ictal measures. Fourth, patients with dual diagnoses and those with indeterminate diagnoses were excluded. Although this strengthened diagnostic separation, it also limits applicability to one of the most clinically challenging groups. Fifth, some patients who answered affirmatively did not provide further elaboration, so misclassification of ordinary postictal relief (eg. relief that the seizure is over) as paradoxical relief cannot be fully excluded. Finally, the mediation analysis is observational and cross-sectional, so the proposed relationship between dissociation, seizure type, and relief should be interpreted as hypothesis-generating rather than causal.

## Conclusion

In conclusion, paradoxical relief was substantially more common after functional/dissociative than epileptic seizures and provided a clinically meaningful, although not definitive, diagnostic signal. Exploratory analyses suggest that pre-ictal dissociative experiences may contribute to the emergence of relief. Prospective, externally validated studies are now required to establish the reproducibility and incremental diagnostic value of this observation, including in patients with dual diagnoses.

## Data Availability

All data produced in the present study are available upon reasonable request to the authors.

## Appendix 1

### Pre-Seizure State

(0) Never, (1) Occasionally, (2) Sometimes, (3) Usually, (4) Always

Before you have your seizure …

Autonomic/Somatic Symptoms

1. Do you feel a shortness of breath or smothering sensation? ⃞
2. Do you have a racing/pounding heart or an increased heart rate? ⃞
3. Do you feel like you’re sweating or have a clammy feeling? ⃞
4. Do you get nausea or a feeling of sickness? ⃞
5. Do you get any form of abdominal distress, such as butterflies or a knot in your stomach? ⃞
6. Do you notice if you get a dry throat or mouth? ⃞
7. Do you get any trembling or shaking of any part of the body? ⃞
8. Do you notice any numbness or tingling sensation in any part of your body? ⃞
9. Is there any head pain or discomfort that you are aware of, like a migraine or pressure? ⃞
10. Is there any pain you feel in any other part of your body, whether sharp or aching character? ⃞
11. Do you experience any change in temperature on your body (e.g., sensation of cold or warmth)? ⃞

Dissociative Symptoms (Depersonalisation, Derealization)

1. Out of the blue, do you feel strange, as if you were not real or were cut off from the world? ⃞
2. Does what you see around you look ‘flat’ or ‘lifeless’, as if you were looking at a picture? ⃞
3. Do parts of your body feel as if they didn’t belong to you? ⃞
4. Do you have the feeling of being a ‘detached observer’ of yourself? ⃞
5. Does your body feel very light, as if it were floating on air? ⃞
6. Do you hear familiar voices (including your own) and do they sound far away and unreal, as if you’re underwater? ⃞
7. Do you have the feeling that your hands or feet have become larger or smaller? ⃞
8. Do your surroundings feel detached or unreal, as if there were a veil between you and the outside world? ⃞
9. Do you have ‘visions’ in which you can see yourself from the outside, as if you were looking at your image in a mirror? ⃞
10. Do you feel that objects around you seem to look smaller or further away? ⃞
11. Do you have the feeling of being outside your body? ⃞

